# Determinants, strategies, and outcomes of implementing an enhanced dosimetry quality assurance checklist in radiation oncology: A qualitative implementation science study

**DOI:** 10.64898/2026.08.05.26359837

**Authors:** Karthik Adapa, Prithima Reddy Mosaly, Fei Yu, Carlton Moore, Ross McGurk, Shiva Das, Lukasz Mazur

## Abstract

Radiation oncology has a long history of developing in-house health information technology (HIT) tools such as quality assurance (QA) checklists, yet there is little guidance from professional bodies on how to implement these tools in complex clinical environments. Building on our previous work that used human-centered participatory co-design, the Task-User-Representation-Function (TURF) framework, and multi-method usability evaluations to design and develop an enhanced dosimetry QA checklist (DQC), this study investigated the barriers and facilitators (determinants) to implementing the enhanced DQC in a radiation oncology clinic, examined implementation strategies, proposed an implementation framework for QA checklists in radiation oncology, and assessed four implementation outcomes: acceptability, appropriateness, feasibility, and adoption. We conducted a qualitative implementation study using an abductive research approach at an academic medical center. All key stakeholders (dosimetrists, physicists, trainees, and software developers) participated in semi-structured interviews, field observations, and surveys across pre-implementation, implementation, and post-implementation phases. Data were analyzed using a hybrid inductive-deductive approach, with deductive coding guided by an adapted Consolidated Framework for Implementation Research (CFIR) mapped to the Unified Theory of Acceptance and Use of Technology and by the Expert Recommendations for Implementing Change (ERIC) compilation. We identified 4 CFIR constructs and 12 sub-constructs as barriers, with structural characteristics and planning showing the highest negative valence, and 5 CFIR constructs and 19 sub-constructs as facilitators, with relative advantage, culture, and leadership engagement showing the highest positive valence. Participants’ suggestions mapped to 19 ERIC strategies in 7 clusters, and the CFIR-ERIC matching tool identified 14 evidence-based strategies in 4 clusters that informed a proposed phased implementation framework. Acceptability, appropriateness, and feasibility scores improved significantly from pre-implementation to implementation for all professional roles (p<0.05), yet adoption reached 100% only in the sixth week of implementation. These findings highlight the value of combining subjective and objective implementation outcomes and provide a practical, evidence-based framework for implementing in-house QA checklists in radiation oncology that warrants validation in diverse settings.

## Introduction

Maximizing patient safety is a central challenge in radiation oncology, and incident learning system studies indicate that many patient safety events originate in the treatment planning stage of the radiation therapy care process [1]. To mitigate these risks, radiation oncology has a long history of developing in-house health information technology (HIT) tools for treatment planning, plan evaluation, quality assurance (QA), and workflow improvement [2–4]. However, there is no guidance or recommendation from professional bodies such as the American Association of Physicists in Medicine (AAPM) or the American Society for Radiation Oncology (ASTRO) on how to develop, implement, and refine these in-house HIT tools [4–6]. Thus, despite their potential, several barriers (e.g., lack of interoperability, information governance uncertainty, and organizational resistance) impede the successful implementation of in-house HIT tools, such as QA checklists, in large and complex healthcare systems [6].

This study is the final stage of a multi-year program of research on dosimetry QA checklists (DQCs) at our institution. In earlier work, we evaluated the perceived usability of our current DQC and explored its association with the perceived cognitive workload of dosimetrists during routine QA tasks in clinical settings [7]. We then used human-centered participatory co-design with key clinical stakeholders to generate design requirements for an enhanced DQC [8], applied the Task-User-Representation-Function (TURF) framework to design the enhanced DQC [9], and evaluated the usability of the DQC and its associations with workload, performance, and patient safety in clinical settings [10]. The enhanced DQC integrates patient and treatment plan information from the two principal clinical information systems used in our clinic (the treatment planning system, RayStation, and the oncology information system, Mosaiq) into a single checklist-driven QA report. The transition of such a tool from laboratory evaluation to routine clinical use, however, involves numerous barriers and challenges that design and usability studies alone cannot address.

Implementation science is “the scientific study of methods to promote the systematic uptake of research findings and other evidence-based practices into routine practice, and, hence, to improve the quality and effectiveness of health services” [11]. Traditionally, implementation science focused on implementing public health interventions, with limited emphasis on implementing HIT tools in healthcare settings [12,13]. Increasingly, however, there is a need to understand how HIT implementation strategies work in complex healthcare systems, for whom, under what determinant conditions, and on what implementation and clinical outcomes [14–16]. There is also limited evidence and guidance regarding the successful integration of HIT tools such as QA checklists into specific clinical settings. Guidelines, theoretical frameworks, models, and recommendations from this evolving discipline can therefore provide insights for the successful and sustainable implementation of in-house HIT tools in radiation oncology clinics.

The aims of this study were: (i) to investigate the barriers and facilitators for implementing the enhanced DQC in a radiation oncology clinic; (ii) to examine the strategies for implementing the enhanced DQC in the radiation oncology clinic; (iii) to propose a framework for implementing QA checklists in radiation oncology; and (iv) to assess the implementation outcomes, specifically acceptability, feasibility, appropriateness, and adoption, of the enhanced DQC in the clinic.

## Materials and methods

### Theoretical framework

In implementation science, there are many models, frameworks, and theories for specific core aspects of implementation research, such as determinants, strategies, mechanisms of action, and outcomes [17–19]. Multiple logic models are available to provide an overarching and coherent rationale for how these core aspects are selected and evaluated with one another [20,21]. Among these, the Implementation Research Logic Model (IRLM) [22] provides a conceptual linkage between the core elements (determinants, strategies, mechanisms of action, and outcomes) and imparts a good understanding of the connections between them when implementing HIT tools such as the enhanced DQC in clinical settings. The IRLM is a semi-structured, principle-guided tool that improves specification, rigor, reproducibility, and transparency in describing the core aspects of implementation projects, and it applies equally to projects in the planning, executing, reporting, and synthesizing stages of research [22]. We therefore used the IRLM as the overarching framework to report on whether and how we accomplished the aims of this study (Fig 1).

**Fig 1.**
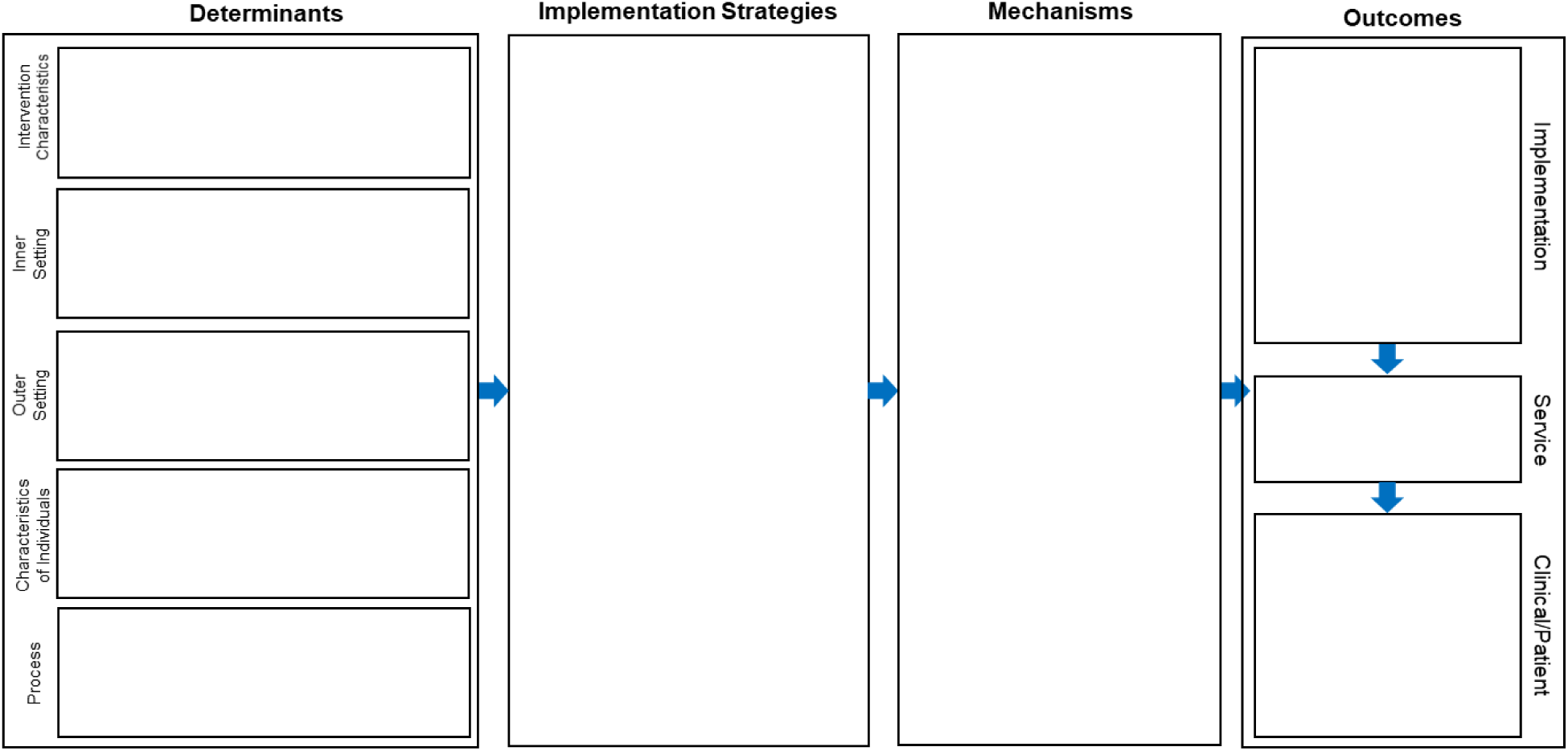
The Implementation Research Logic Model (IRLM). Core elements of the IRLM linking determinants, implementation strategies, mechanisms of action, and outcomes. Adapted from Smith, Li, and Rafferty (2020) [22], distributed under the terms of the Creative Commons Attribution 4.0 International License.

#### Determinants

Determinants are factors that might prevent or enable implementation (i.e., barriers and facilitators); they may act as moderators or mediators and serve as links in a chain of causal mechanisms [23]. Common determinant frameworks are the Consolidated Framework for Implementation Research (CFIR) [24–26] and the Theoretical Domains Framework (TDF). Between these two widely used frameworks, the CFIR, with 39 constructs, comprehensively covers potential determinants for both implementation and effectiveness within a healthcare setting, while the TDF, with 12 constructs, focuses on a coherent explanation of providers’ behavioral change [27]. The CFIR enables the description of factors related to both the general implementation context and intervention-specific factors [28,29]. Hence, we used the CFIR to identify determinants of the enhanced DQC’s implementation. The CFIR constructs are organized into five domains: intervention characteristics (e.g., complexity, strength of the evidence), outer setting (e.g., external policy and incentives), inner setting (e.g., organizational culture, the extent to which leaders are engaged), characteristics of individuals (e.g., self-efficacy), and process (e.g., planning and engaging key stakeholders) [25,28,30]. This information can guide decisions about the types of strategies that may be appropriate and match the needs of the context.

Studies suggest that the CFIR must be adapted because the framework was largely aimed at implementing singular, face-to-face intervention-based services rather than HIT tools [29]. We therefore adapted the CFIR to the enhanced DQC based on guidelines from previous studies [19,31] (for example, the intervention characteristics construct was adapted as QA checklist characteristics, intervention source was adapted as source of the QA checklist, and patient needs and resources was adapted as user needs and resources). Recent studies focused on implementing HIT tools suggest that existing implementation science frameworks do not completely account for the complexities of healthcare systems; we therefore mapped the considerations proposed in Marwaha and colleagues’ framework for deploying digital health tools within large, complex health systems [32] to the adapted CFIR. Further, because implementing the enhanced DQC requires attention to factors related to both practice change and technology acceptance, we also mapped the constructs of the Unified Theory of Acceptance and Use of Technology (UTAUT) [33,34], one of the best developed and most widely used technology acceptance theories, to the adapted CFIR (S1 Table). Demographic constructs of the UTAUT (age, gender, and professional experience) were not considered because of the limited participant size. The adapted CFIR constructs and an example interview question for each CFIR sub-construct are provided in S1 Appendix.

#### Implementation strategies

Implementation strategies refer to supports, changes to, and interventions on the system to increase the adoption of the enhanced DQC in the clinic. Determinants are commonly used to select and tailor implementation strategies; however, because success or failure is often determined by multiple factors within a dynamic delivery system, the field has experienced challenges identifying consistent links between individual barriers and specific strategies to overcome them. The Expert Recommendations for Implementing Change (ERIC) compilation is a comprehensive compilation of 73 discrete strategies to facilitate implementation in clinical practice [35], which have been grouped into 9 clusters based on concept mapping [36]. CFIR domains and ERIC strategies are coherent and synergistic, and a CFIR-ERIC matching tool has been developed to determine which strategies would best address contextual barriers identified by the CFIR [36,37]. The tool was developed by asking implementation science experts to rank the top seven ERIC strategies that, in their view, would address barriers categorized by each CFIR construct; it allows users to indicate which determinants are relevant and generates an output table matching CFIR constructs to ERIC strategies with the percentage of experts endorsing each strategy for each construct [30,37,38]. We used the CFIR-ERIC matching tool because it provides a structured approach to considering a broad range of strategies for the identified barriers and facilitators.

#### Mechanisms of action

Mechanisms of action refer to “processes or events through which an implementation strategy operates to affect desired implementation outcomes” [23]. A mechanism may be a change in a determinant, an early implementation outcome, an implementation strategy, or a combination of these factors [39]. Few implementation studies have formally tested mechanisms of action, although attention to this area is increasing [23]. We did not consider this element of the IRLM in this study because we could not evaluate all implementation outcomes.

#### Implementation outcomes

Implementation outcomes are the “effects of deliberate and purposive actions to implement new treatments, practices, and services” [40–42]. They may be indicators of implementation processes, key intermediate outcomes concerning services, or target clinical outcomes. RE-AIM (Reach, Effectiveness, Adoption, Implementation, and Maintenance) and Proctor and colleagues’ implementation outcomes framework (IOF) are the most widely used outcomes frameworks in dissemination and implementation research [12,43]. The RE-AIM framework includes six dimensions and focuses on outcomes at the individual and organizational levels [44]. The IOF presents eight implementation outcomes (acceptability, appropriateness, cost, fidelity, feasibility, penetration, adoption, and sustainability) [42,43], distinguishes between implementation, service, and patient outcomes, and examines relationships across implementation outcomes at various stages of implementation research [42,45,46]. We therefore used the IOF to assess implementation outcomes. Recent reviews of implementation outcomes research have highlighted the lack of validated instruments to measure outcomes such as penetration, fidelity, and sustainability for HIT tools [45,47]. In this study, we used the IOF to assess four key outcomes (acceptability, feasibility, appropriateness, and adoption) of implementing the enhanced DQC in clinical settings. The remaining implementation outcomes (cost, penetration, fidelity, and sustainability) and service and patient outcomes were not part of this study.

### Study setting

We conducted this study in the radiation oncology clinic of an academic medical center (University of North Carolina at Chapel Hill, United States). The enhanced DQC was implemented in the clinic on March 15, 2022, after an elaborate multi-method development and evaluation program described in our previous publications [8–10]. In this clinic, the enhanced DQC (internally referred to as V3) was piloted in near-live conditions alongside the current DQC (V2) before its clinical release.

### Study design

This is a qualitative implementation study based on an abductive research approach, as described by Dubois and Gadde [48] and Zainal [49]. In this approach, inductive and deductive analysis methods are systematically combined using existing theories, and the results are further refined to match the specific context (a QA checklist in a radiation oncology clinic). We considered this approach appropriate for deriving practical recommendations for implementing an enhanced QA checklist in the radiation oncology clinic. Transcripts of interviews and online survey questionnaires with key stakeholders in the clinical implementation process, together with the results of field observations, huddle meetings, research team meetings, and informal discussions within the research group, were analyzed and applied to the CFIR domains and ERIC strategies. Finally, we used the CFIR-ERIC implementation strategy matching tool to identify evidence-based strategies, which were used to develop the proposed implementation framework (Fig 2).

**Fig 2.**
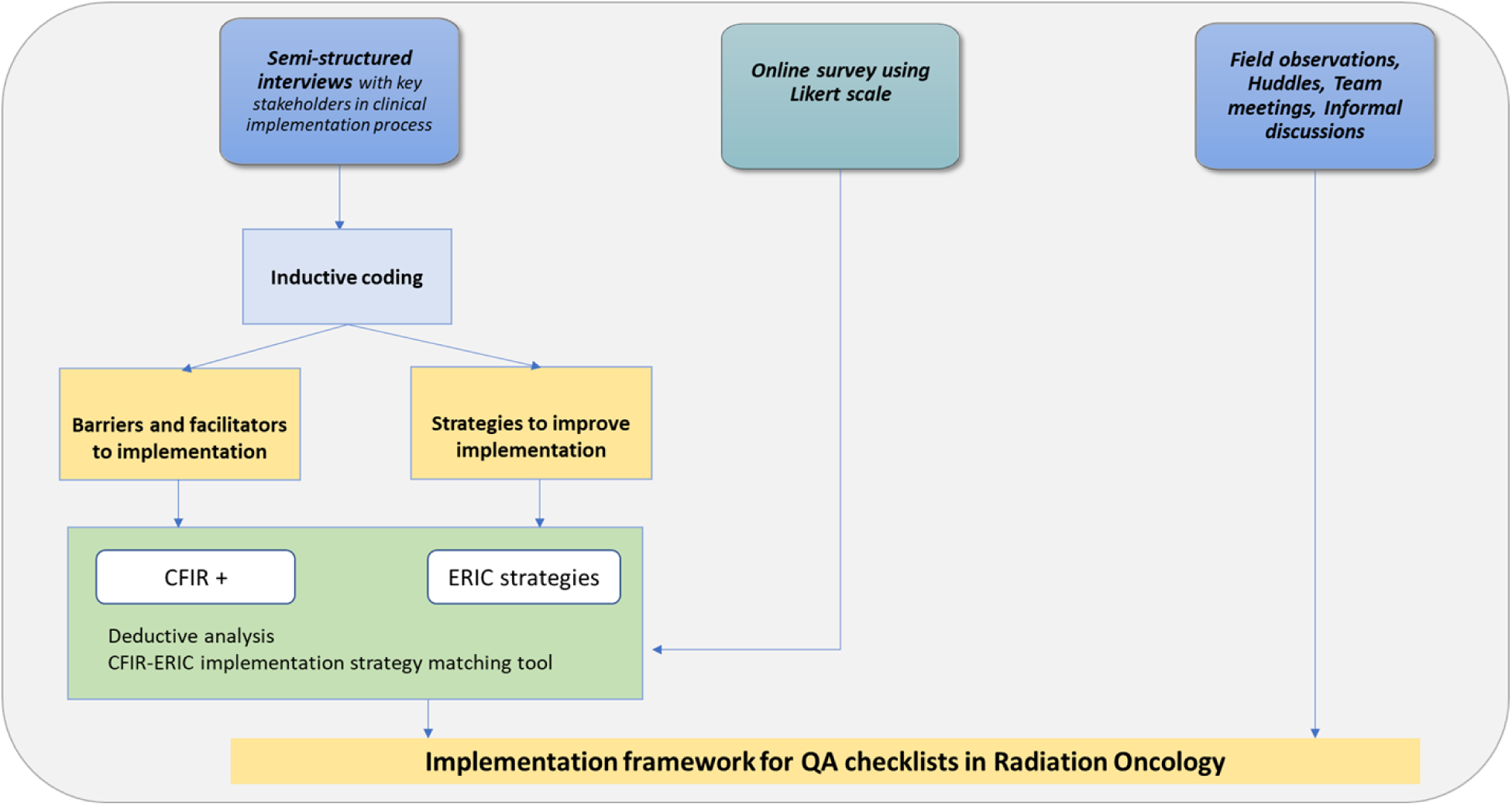
Study design. Overview of the data sources, analysis steps, and outputs of the qualitative implementation study across the pre-implementation, implementation, and post-implementation phases of the enhanced dosimetry quality assurance checklist (DQC).

### Participants

The study participants comprised all key stakeholders involved in implementing the enhanced DQC at the academic medical center: dosimetrists, physicists, trainees, and software developers. We approached all eligible participants and used purposive sampling with the aim of including stakeholders who were actively involved in implementing the enhanced DQC and who represented all professional groups. The identified participants had closely experienced the enhanced DQC’s implementation by overseeing the implementation process, receiving feedback about the system from other care team members, and using the enhanced DQC in their clinical practice.

### Data collection

Table 1 provides a summary of the data collection methods and the number of participants for each method across the three stages of the enhanced DQC’s implementation: pre-implementation (January 31 to March 14, 2022), implementation (March 15 to April 30, 2022), and post-implementation (July 15 to August 30, 2022).

**Table 1.** Summary of data collection methods and number of participants by implementation phase.

| Data collection method | Pre: D (n=5) | Pre: P (n=11) | Pre: T (n=4) | Impl: D (n=5) | Impl: P (n=11) | Impl: T (n=4) | Post: D (n=6) | Post: P (n=11) | Post: T (n=4) | Post: Dev (n=2) |
| --- | --- | --- | --- | --- | --- | --- | --- | --- | --- | --- |
| Semi-structured interviews | 3 | 2 | 2 | 4 | 2 | 2 | 5 | 6 | 0 | 2 |
| Observations | 4 | 0 | 0 | 5 | 4 | 2 | 2 | 2 | 0 | 0 |
| Survey for determinants | 0 | 0 | 0 | 0 | 0 | 0 | 4 | 6 | 0 | 2 |
| Survey for outcomes | 4 | 5 | 4 | 4 | 5 | 4 | 0 | 0 | 0 | 0 |

Pre = pre-implementation (January 31–March 14, 2022); Impl = implementation (March 15–April 30, 2022); Post = post-implementation (July 15–August 30, 2022); D = dosimetrists; P = physicists; T = trainees; Dev = software developers.

#### Semi-structured interviews, observations, and surveys

Semi-structured interviews were conducted in person or remotely with dosimetrists, physicists, and trainees during pre-implementation, implementation, and post-implementation. We also interviewed the software developers (2/2, 100%) post-implementation. Pilot interviews with three participants were used to refine the interview outline, and the formal outline was finalized after appropriate modifications. During the interviews, communication techniques such as rhetorical questioning and repetition were used to reflect the views of the interviewees as truthfully and comprehensively as possible. All interviews were recorded with participants’ oral consent; each interview lasted approximately 30 to 60 minutes and was conducted one to two times to ensure data integrity. Sampling continued until saturation was achieved, when new data ceased to be generated. The interview guide was based on the adapted CFIR categories mapped to Marwaha and colleagues’ framework [32] and the UTAUT (S2 Appendix).

We conducted field observations during pre-implementation and from the first through the sixth week of implementation (dosimetrists, n=5; physicists, n=4; trainees, n=2). The observation guide is provided in S3 Appendix. Observations helped us gain a more objective perspective and minimize potential biases that naturally arise when using a qualitative research approach.

We also administered a cross-sectional survey on determinants to 12 participants (dosimetrists, n=4; physicists, n=6; software developers, n=2). The survey included questions on CFIR constructs for key determinants, using a five-point Likert-type ordinal response format with the options *not correct at all*, *not quite correct*, *partly correct*, *quite correct*, and *completely correct*. Survey data were collected and managed using an anonymous survey administered via Qualtrics (S4 Appendix).

#### Implementation outcome measures

Table 2 provides the definition, theoretical basis, salience by implementation stage, measurement, and participants for each implementation outcome. Participants (dosimetrists, physicists, and trainees) rated the acceptability, appropriateness, and feasibility of the enhanced DQC’s implementation on a scale of 1 to 5 (1 = completely disagree; 5 = completely agree) across 12 items (total score of 60; 20 per outcome) using the validated four-item Acceptability of Intervention Measure (AIM), Intervention Appropriateness Measure (IAM), and Feasibility of Intervention Measure (FIM) [50]. These measures have previously shown a correlation with implementation success [42] and have been used in radiation oncology settings [51]. We administered the validated surveys during pre-implementation and during the first two weeks of the enhanced DQC’s implementation. Adoption was assessed objectively as the number of times the enhanced DQC was run compared with the total number of times the enhanced and current DQC were run by all participants per week during the first six weeks of implementation; this metric has been used in previous studies as a measure of HIT tool adoption [52].

**Table 2.** Taxonomy of implementation outcomes assessed in this study (adapted from Proctor et al., 2011 [42]).

| Implementation outcome | Working definition | Theoretical basis | Salience by implementation stage | Measurement | Participants |
| --- | --- | --- | --- | --- | --- |
| Acceptability | The perception among implementation stakeholders that the enhanced DQC is agreeable, palatable, or satisfactory | Rogers: complexity and, to a certain extent, relative advantage | Early | Validated survey (AIM) administered pre-implementation and during early implementation | Dosimetrists, physicists, trainees |
| Appropriateness | The perceived fit and relevance of the enhanced DQC | Rogers: compatibility | Early (before adoption) | Validated survey (IAM) administered pre-implementation and during early implementation | Dosimetrists, physicists, trainees |
| Feasibility | The extent to which the enhanced DQC can be successfully used in the radiation oncology clinic | Rogers: compatibility and trialability | Early (during adoption) | Validated survey (FIM) administered pre-implementation and during early implementation | Dosimetrists, physicists, trainees |
| Adoption | The extent to which the enhanced DQC has been employed by key stakeholders | RE-AIM: adoption; Rogers: trialability (particularly for early adopters) | Early to mid | Number of enhanced DQC runs divided by the total number of enhanced and current DQC runs per week during the first six weeks of implementation [52] | Dosimetrists, physicists, trainees |

DQC = dosimetry quality assurance checklist; AIM = Acceptability of Intervention Measure; IAM = Intervention Appropriateness Measure; FIM = Feasibility of Intervention Measure.

### Data analysis

We applied a hybrid approach combining inductive and deductive coding elements, as described by Fereday and Muir-Cochrane [53]. This approach incorporates both the data-driven inductive approach of Boyatzis [54] and the deductive a priori template of codes approach outlined by Crabtree and Miller. The hybrid approach helps unearth how participants make sense of, or interpret, the implementation of QA checklists (inductive analysis) while also providing the “ideal types” from the CFIR and ERIC frameworks to interpret and describe the implementation of the enhanced DQC in the clinic.

First, we analyzed the semi-structured interview transcripts using a thematic analysis approach with an inductive coding process. Themes and codes were iteratively developed and applied to all transcripts, and the content of the codes was summarized to obtain the main findings used for the deductive analysis [54]. For the deductive analysis, we used two templates: the CFIR domains and the ERIC strategies grouped into 9 clusters [36]. The first author coded the summaries from the inductive analysis and the survey findings according to the CFIR domains and ERIC strategies; survey data were therefore not analyzed using quantitative methods. Specifically, the CFIR template was used to analyze the summaries regarding implementation performance, whereas the ERIC strategies served as a template for analyzing the summaries of participants’ suggestions for improving the enhanced QA checklist’s implementation process. All coding was performed using NVivo qualitative data analysis software. Valence for determinants (barriers or facilitators) was determined by the influence that the coded data had on the implementation process, that is, whether contextual factors facilitated or hindered the implementation. When some comments for a determinant were negative and others positive, a mixed (M) rating was applied.

The implementation framework for QA checklists in radiation oncology was derived from the results of the CFIR– and ERIC-guided analyses. The CFIR-ERIC implementation strategy matching tool supported the prioritization of the derived recommendations [37,55]. Findings from the field observations, huddles, team meetings, and informal meetings were used to organize the suggestions provided by the participants for QA checklist implementation. This methodology has been used in previous studies [56,57].

Acceptability, feasibility, and appropriateness scores by role were compared between pre-implementation and implementation using the Wilcoxon signed-rank test. Adoption rates for the enhanced DQC and the current DQC by role were calculated as the percentage of enhanced DQC runs divided by the total number of enhanced and current DQC runs. The adoption of the enhanced DQC was compared with the current DQC using the Mann-Whitney U test. Statistical significance was set at p<0.05.

### Ethics statement

The Institutional Review Board of the University of North Carolina at Chapel Hill reviewed and approved this study [IRB # 20-1355. All interviews were recorded with participants’ oral informed consent, and survey responses were collected anonymously.

## Results

### Overview

Inductive analysis of the interview transcripts revealed two broad categories: the implementation process of the enhanced DQC and participants’ suggestions for improving the enhanced DQC’s implementation in the clinic. Three themes emerged regarding the implementation process: QA checklist features, IT infrastructure and user training, and engaging users. The suggestions provided by interviewees were organized into two themes: involvement of users, and user training and support. Deductive coding revealed 4 CFIR constructs and 12 sub-constructs as barriers, and 5 CFIR constructs and 19 sub-constructs as facilitators. The suggestions for improving the enhanced DQC’s implementation were mapped onto 7 clusters and 19 ERIC strategies.

### Determinants

#### Implementation process

##### QA checklist features

Interviewees highlighted that the enhanced QA checklist presents information from both Mosaiq and RayStation and is a major improvement over the current DQC. However, they also worried that they might sometimes rely completely on the checklist without actually verifying the information in both information systems. They reported that during near-live testing, far too many changes were made rapidly, and they could not completely track all the key changes made each week; some participants noted that a high-level overview of the key changes demonstrated during huddles was a good starting point for key conversations. There were contradictory opinions among users on the design and layout of the enhanced QA checklist. One group of participants highlighted that the enhanced DQC’s color coding, font, and overall design resemble Mosaiq, and they felt comfortable with that familiarity because they had used Mosaiq for a long time (more than a decade for some users). Other users preferred a brighter, newer design and layout completely different from Mosaiq. There were also differing views on the extent of information provided: some interviewees wanted only summary or key information, keeping the interface less busy and using flags to identify errors, while others suggested providing all requisite information and leaving it to the user to decide between “all okay,” “attention,” or “not applicable.”

##### IT infrastructure and user training

Interviewees reported that after the enhanced QA checklist was implemented in the clinic, the tool sometimes could not be run because IT infrastructure problems (e.g., a server without Python installed, a fully loaded disk, or updates to RayStation and Mosaiq) completely derailed the highly sensitive enhanced DQC. Users were extremely frustrated with regular failures and a lack of communication from the developers about the reasons for the crashes, and they highlighted the need for an alternative QA checklist independent of RayStation for future failures. Some participants reported that the enhanced DQC was easy to use and required minimal training. Newly hired employees and trainees reported that one-on-one training (at-elbow support) on the key features of the enhanced DQC, together with sharing best practices for using it, would be more useful than user manuals.

##### Engaging users

Interviewees reported that taking part in implementing the enhanced DQC alongside clinical duties was extremely difficult and resulted in delays in planning and executing the enhanced DQC in the clinic. This was further exacerbated by remote working conditions and a lack of protected time to participate in the multiple meetings required for implementation. Participants also highlighted the need to include a physicist or dosimetrist in the final coding process in the final weeks before implementation, because key decisions for refining the DQC were made based on suggestions provided by participants. All participants agreed that the department’s culture of promoting patient safety and developing new HIT tools such as QA checklists enhanced their confidence that the enhanced DQC would be implemented in the clinic. Some interviewees also highlighted resistance among some users to changing their clinical workflow; these users saw the enhanced DQC as a tool that “standardizes” workflow and does not promote individual freedom to choose the QA checks most appropriate for each plan.

#### Mapping of CFIR domains: barriers

The summaries of participants’ interview transcripts and survey responses were coded and assigned to CFIR domains, and mapped as barriers or facilitators based on valence (a negative influence on implementation was classified as a barrier, and a positive influence as a facilitator). We identified 4 CFIR constructs and 12 sub-constructs as barriers (Table 3). Of these 12 sub-constructs, 5 had both positive and negative comments and were therefore rated as mixed (M). Structural characteristics and planning had the highest negative valence among the 12 CFIR sub-constructs.

**Table 3.**
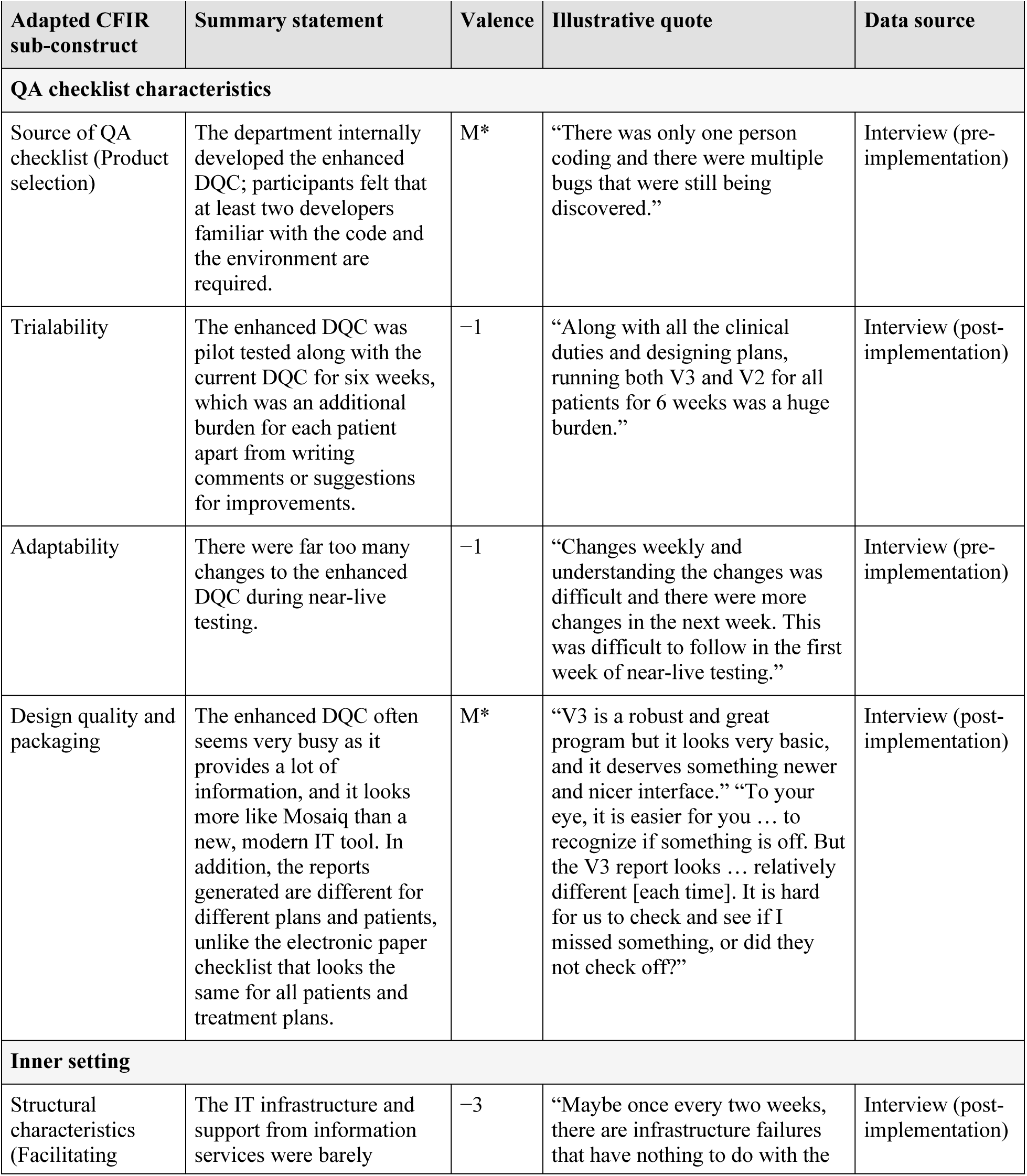
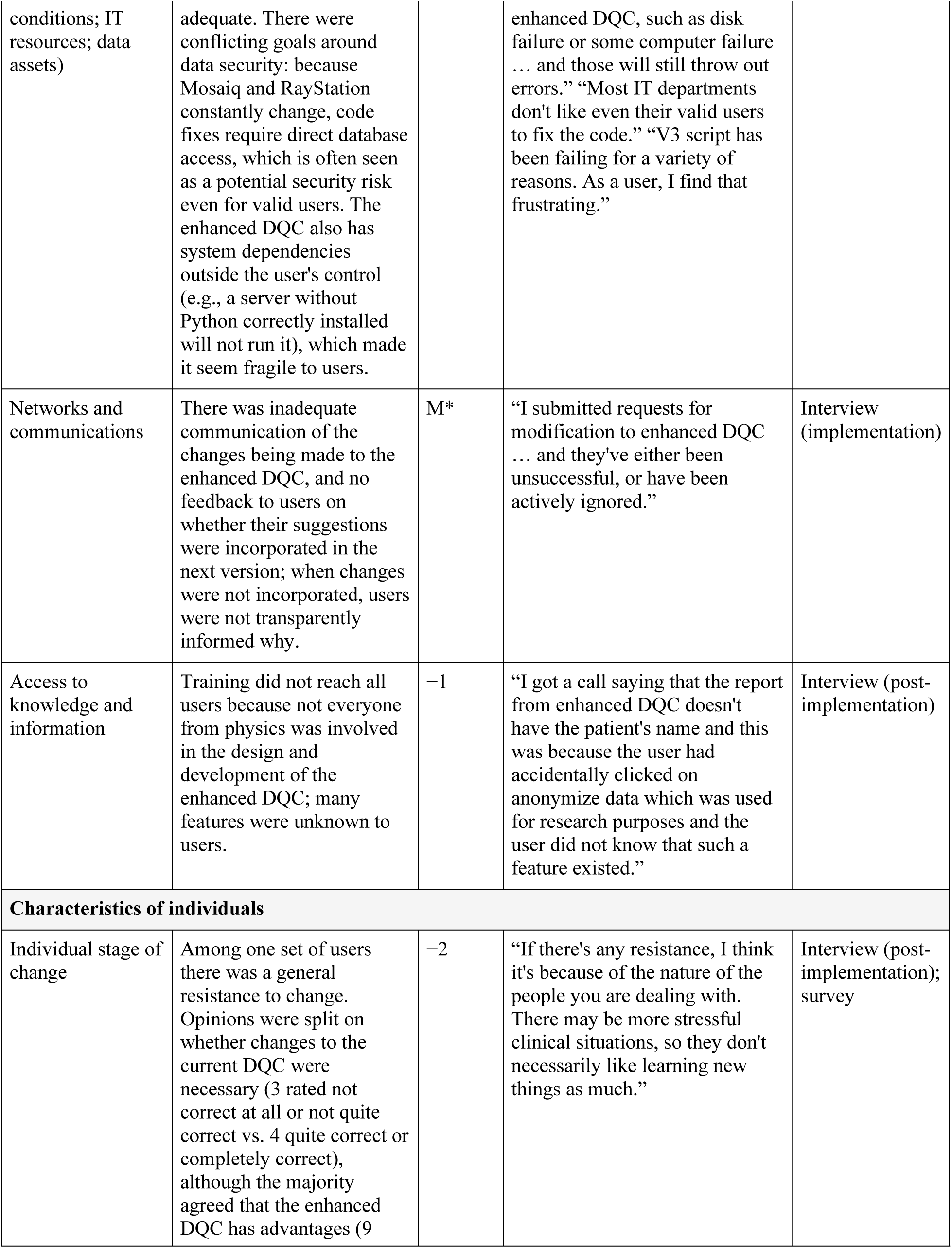
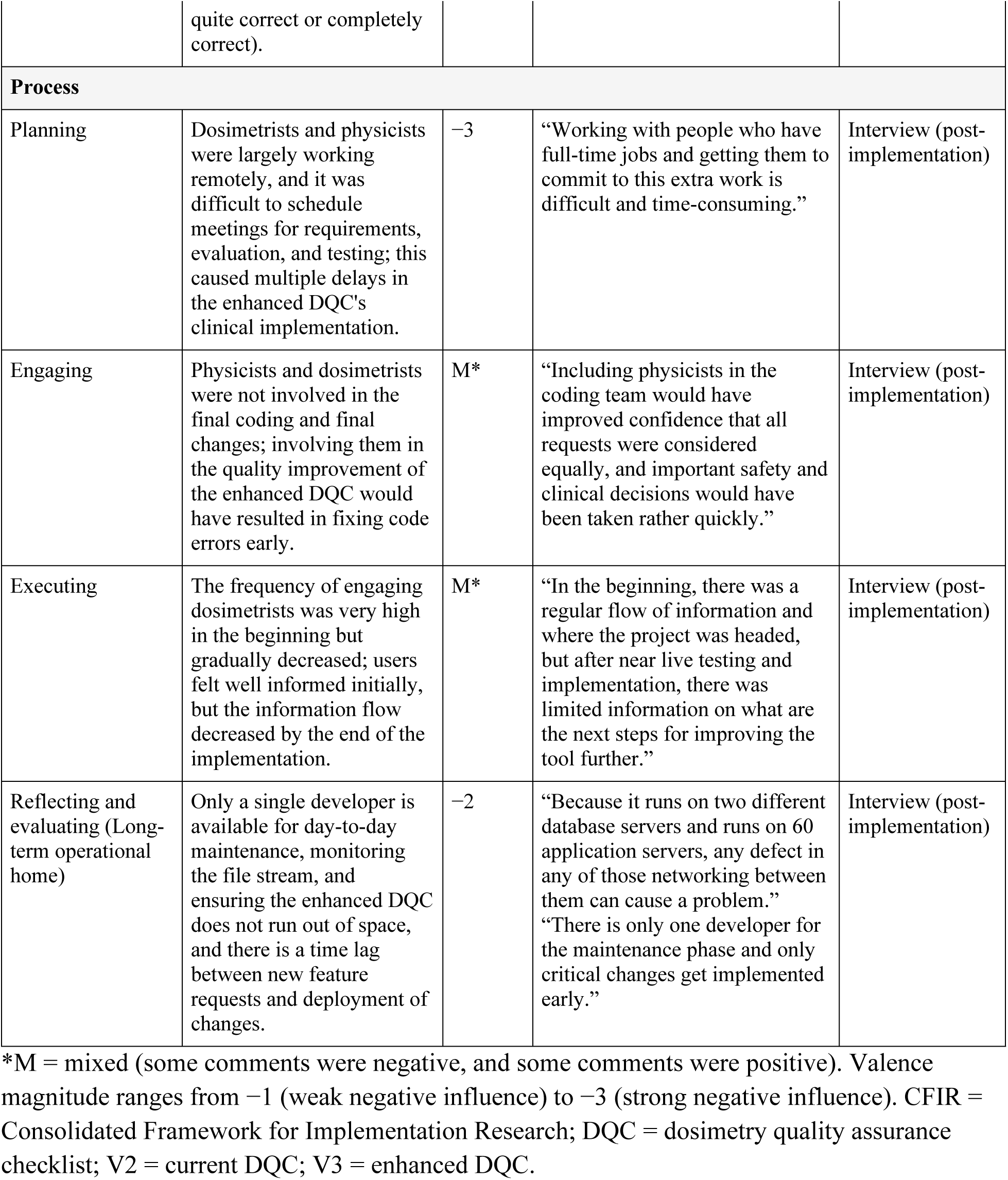
Barriers to implementing the enhanced QA checklist: codes mapped to adapted CFIR domains.

#### Mapping of CFIR domains: facilitators

Summary statements with positive valence were mapped to the CFIR and categorized as facilitators of the enhanced DQC’s implementation in the clinic. We mapped 5 CFIR constructs and 19 sub-constructs as facilitators (Table 4). Of these 19 sub-constructs, 5 were rated as mixed. Relative advantage, culture, and leadership engagement were rated with the highest valence (+4) among the 19 facilitators.

**Table 4.**
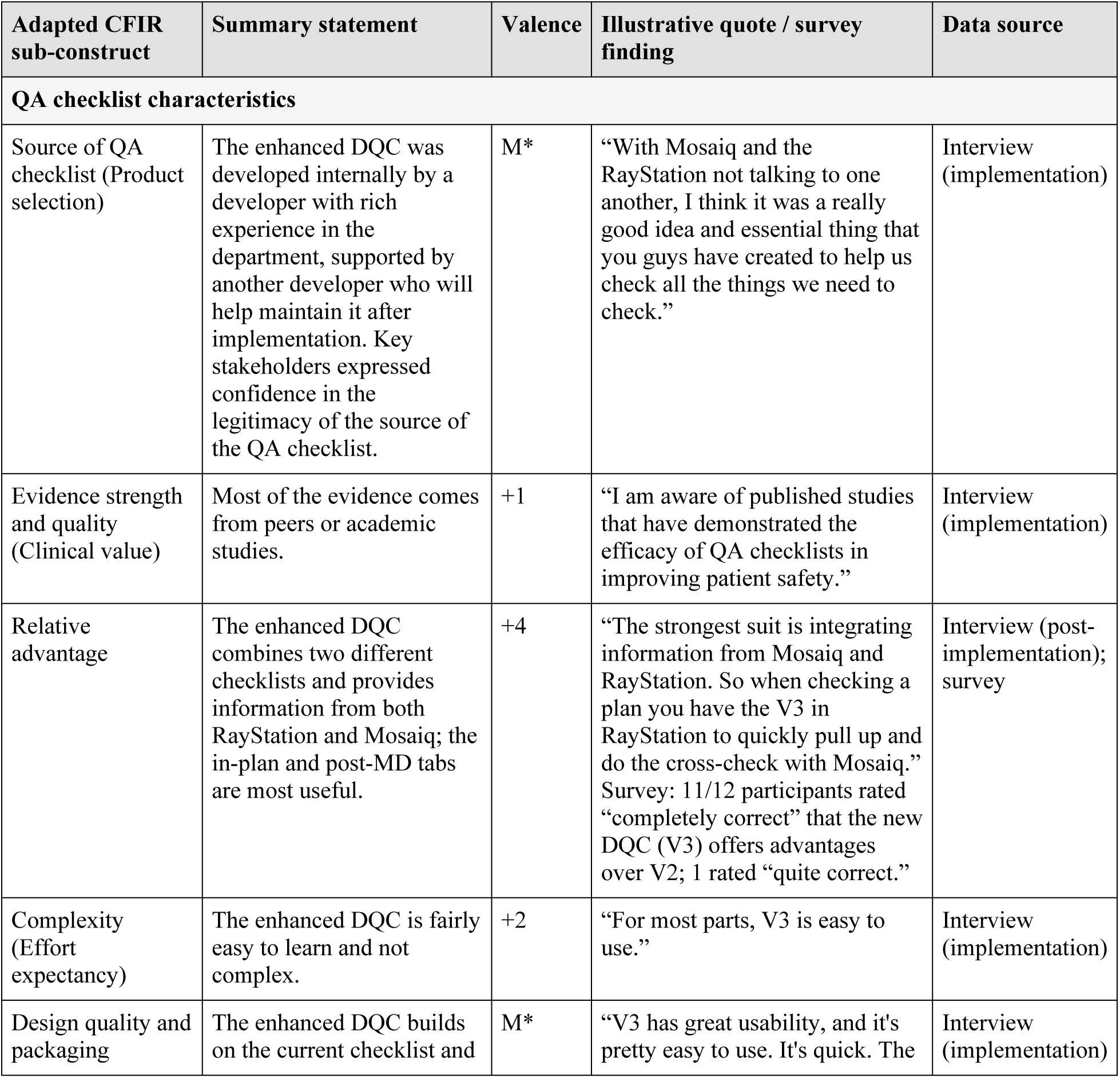
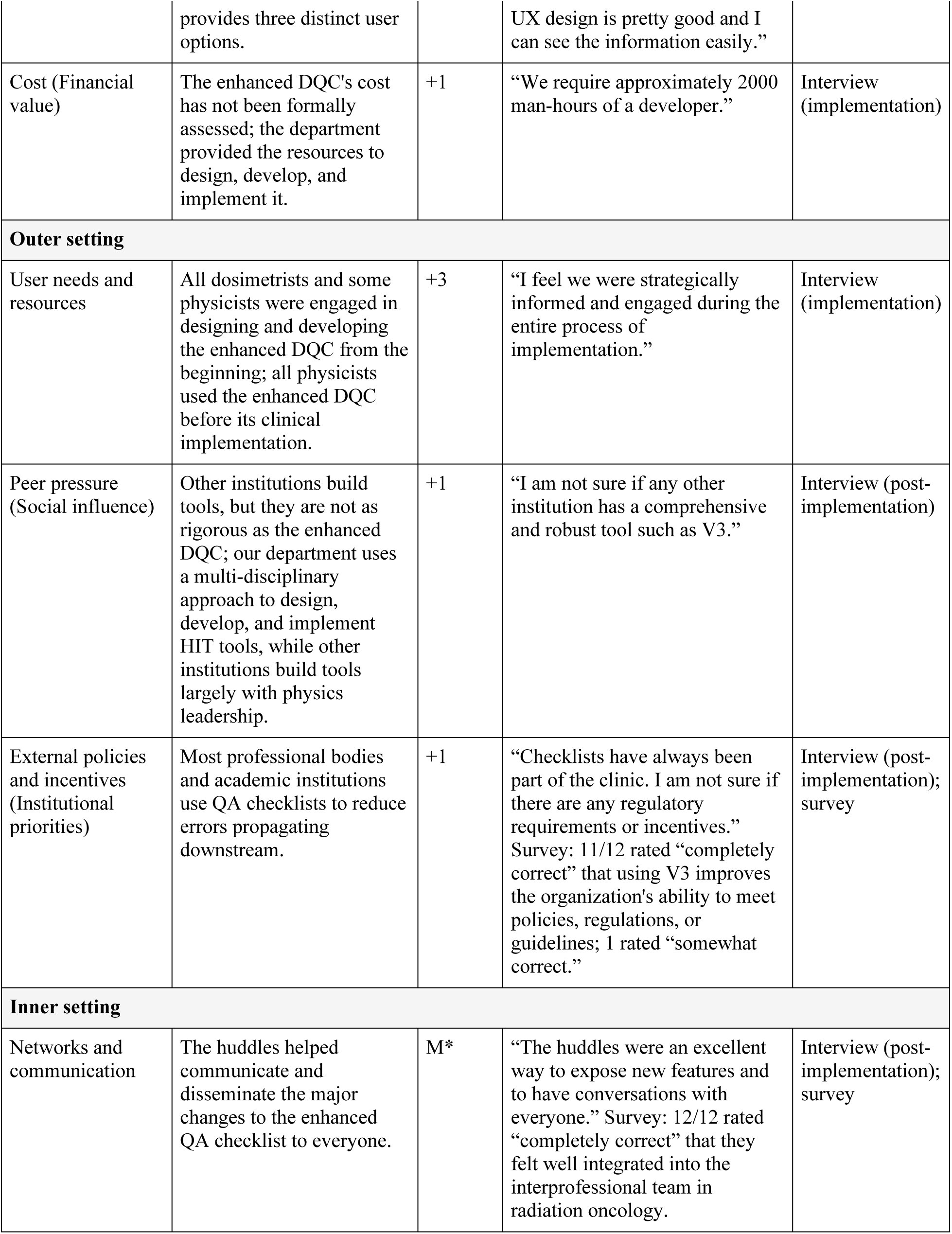
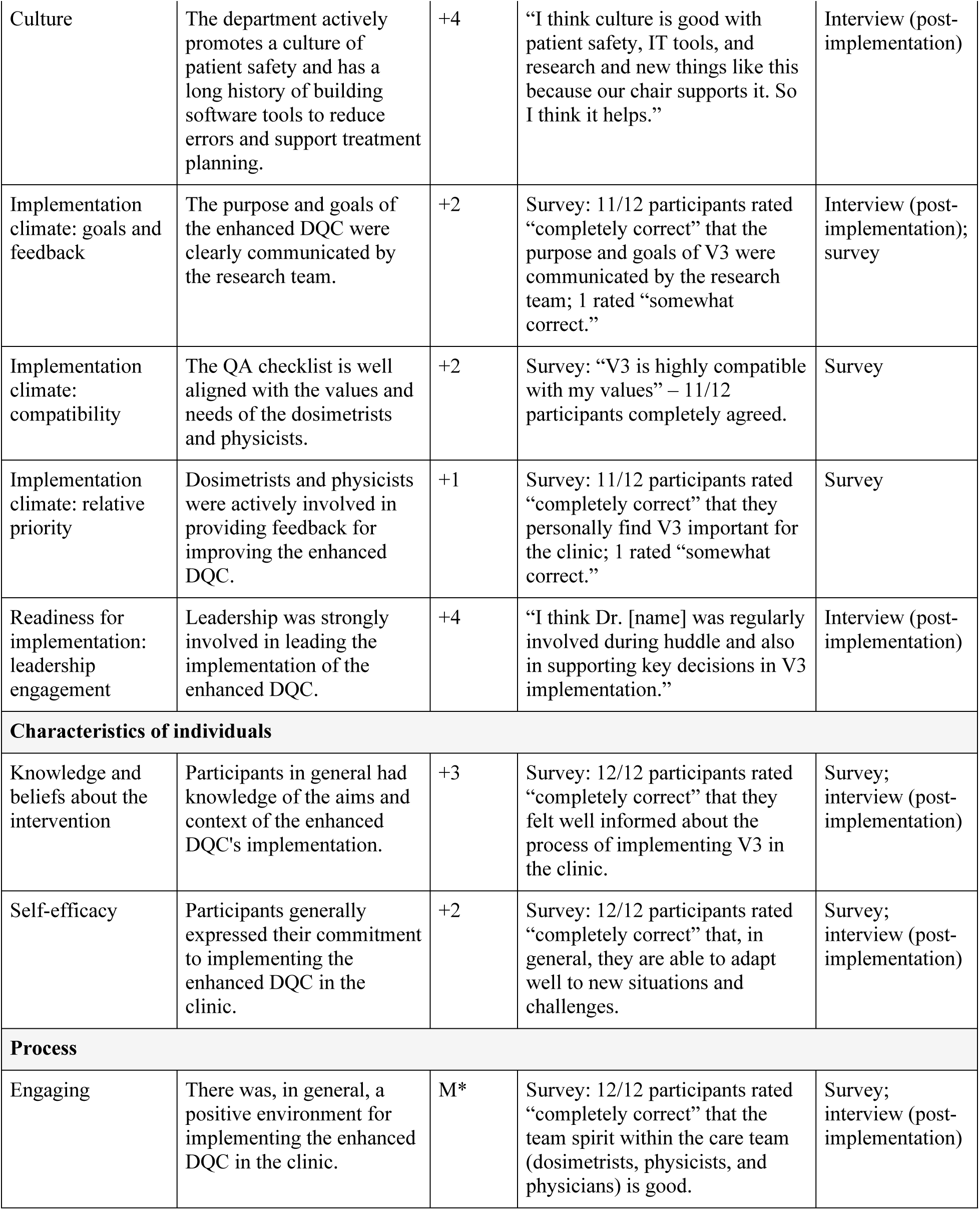
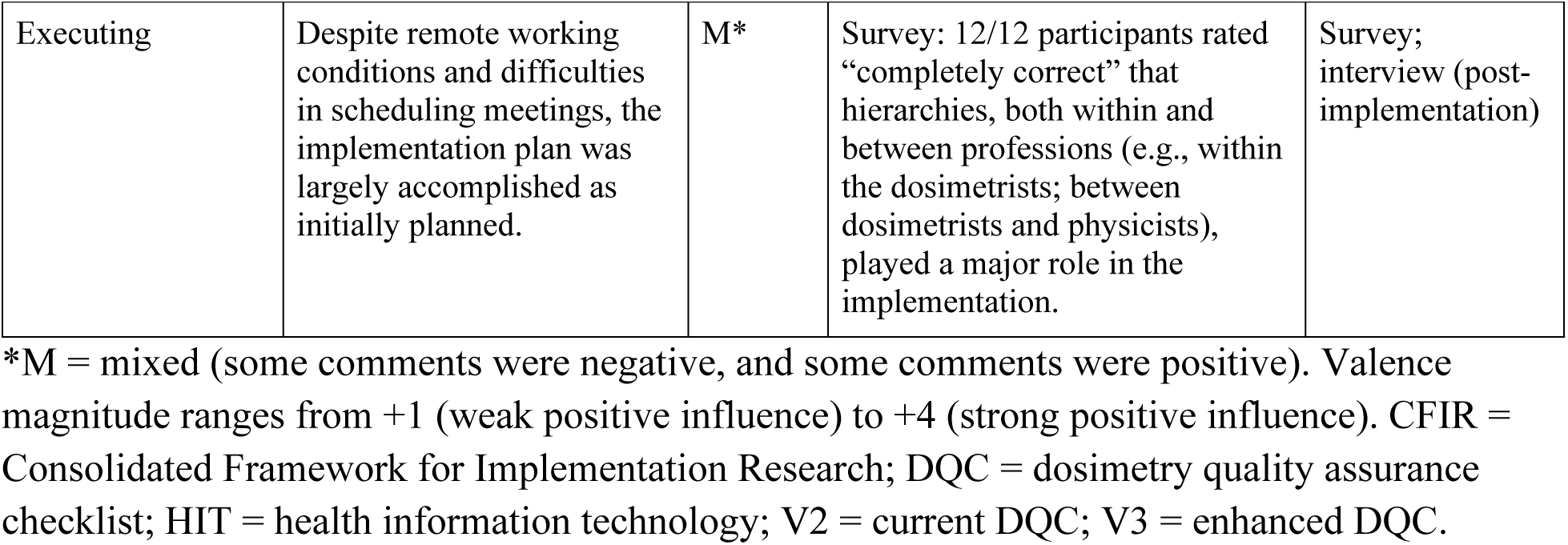
Facilitators of the enhanced QA checklist implementation mapped to CFIR domains.

### Strategies to improve implementation

Interviewees’ suggestions to improve the implementation of the enhanced DQC were categorized into two themes.

#### Involvement of users

Users suggested involving dosimetrists and physicists early in implementing the enhanced DQC, creating a project team that includes at least one physicist and one dosimetrist, and providing them weekly protected time to take part in the first phases of the study. They also noted that the multi-disciplinary team with human factors researchers and developers worked well, but they would prefer a small sub-group of physicists and dosimetrists to conduct a quality check on the tool during its final stages of deployment. Deploying the tool in near-live testing was helpful, but they would prefer testing on a smaller group of users rather than all members of the clinic. They recommended identifying potential users to support and drive the implementation within each group. Users also noted that resolving conflicts or differences of opinion can be challenging, as some user groups are highly collaborative and others less so; they suggested involving a combination of physicists and dosimetrists, rather than dosimetrists alone, so the groups could learn from and work with each other and exchange innovative ideas.

#### User training and support

Most interviewees suggested developing a variety of training materials to match varied user needs: some prefer emails for updates, some are likely to use videos, and most prefer one-on-one training. While overall training needs were limited, interviewees recognized that needs would be higher for new hires and trainees. They recommended that crashes and failures related to the enhanced DQC, irrespective of the cause (internal, such as bugs, or external, such as server breakdowns), be communicated immediately to users. They also recommended including at least two developers in developing the QA checklist and allocating more resources for maintenance and support. They felt that changes were made quickly during near-live testing and implementation, and that post-implementation response times for critical changes were optimal but inordinately slow for routine changes. Users also suggested that, after successful implementation at the medical center, the learnings should be shared and disseminated to satellite sites; some users support radiation oncologists at these sites, which use different processes, so creating a uniform QA process using the enhanced DQC would be a logical next step.

##### Mapping of ERIC strategies

The summary segments of participants’ suggestions for implementation were mapped to the 73 ERIC strategies. We mapped the summary statements to 7 clusters and 19 ERIC strategies, accounting for 26% of all ERIC strategies (Table 5).

**Table 5.**
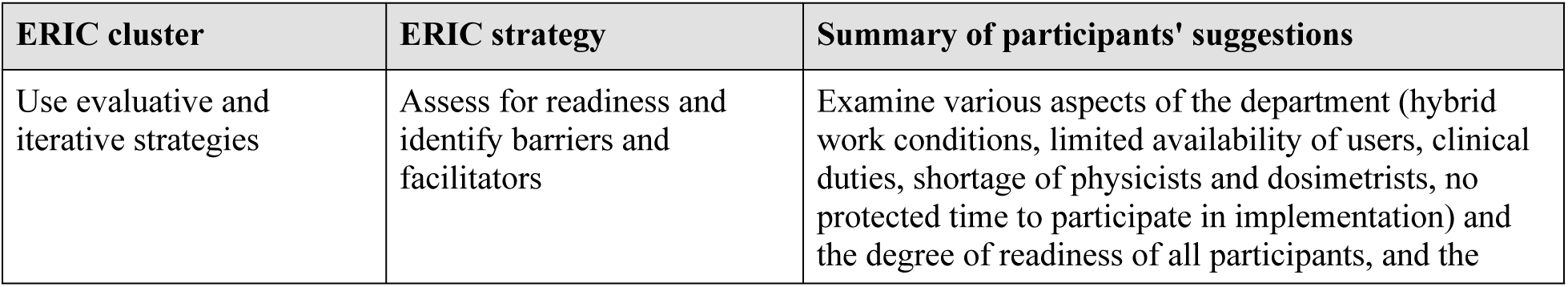
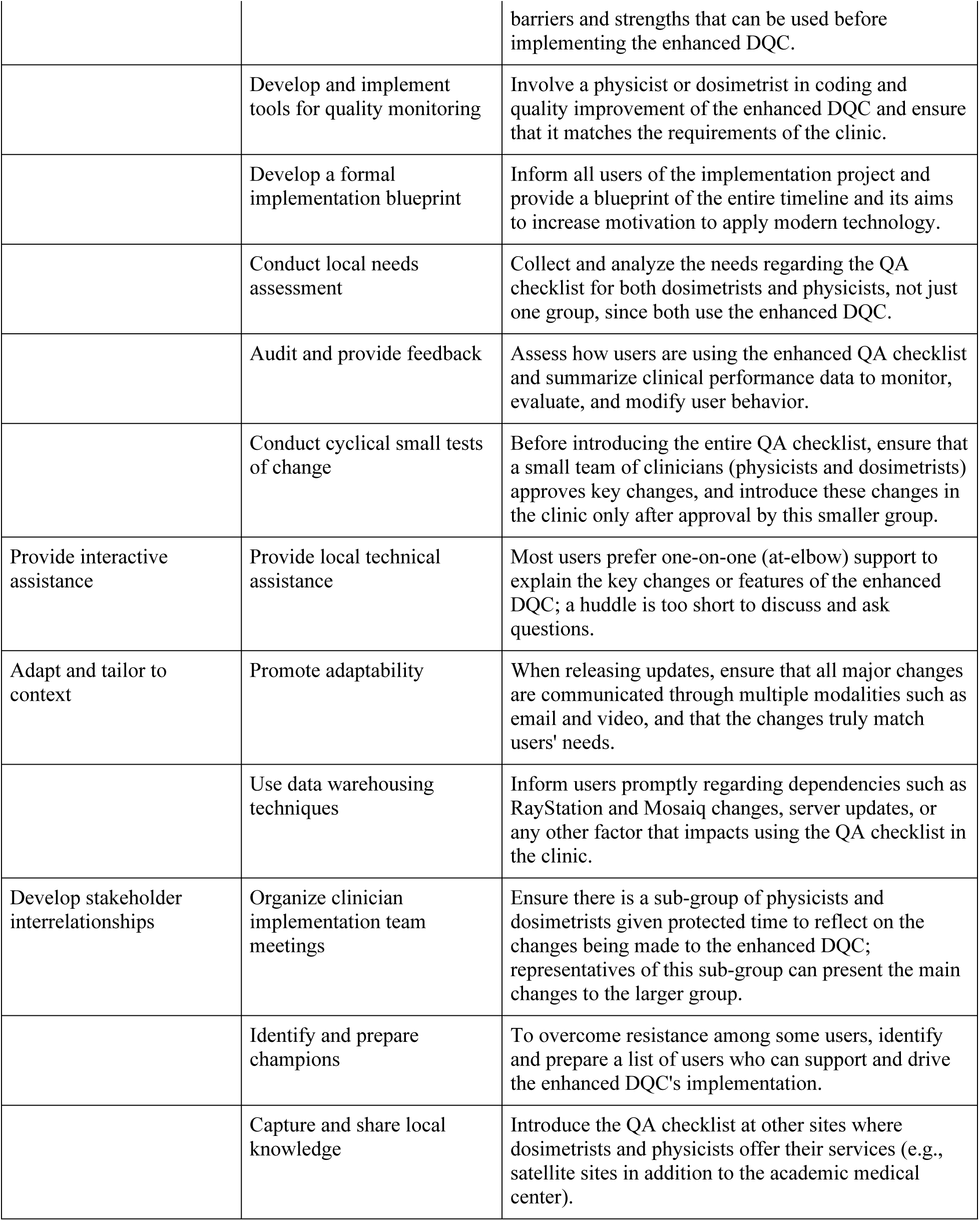
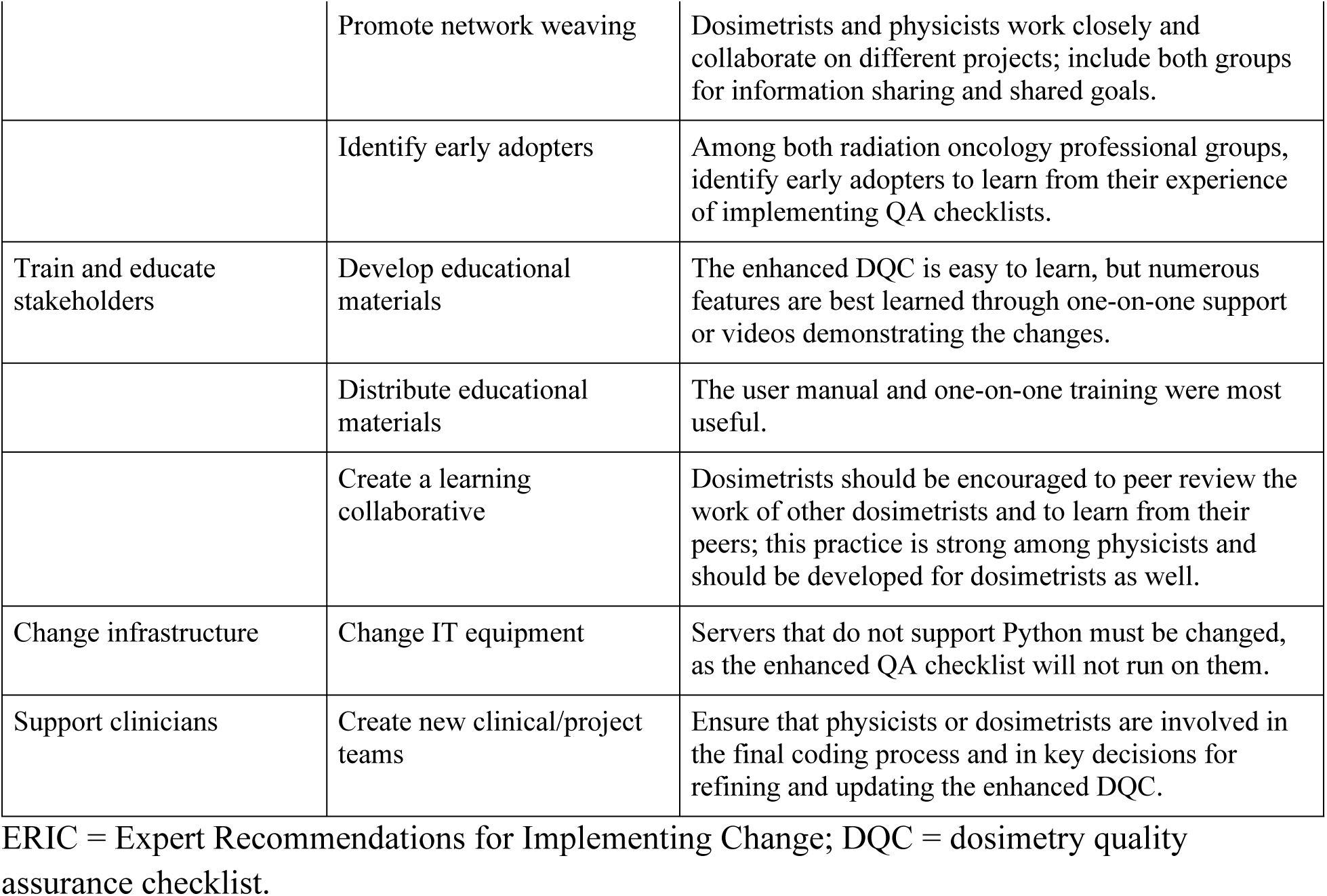
ERIC strategies mapped to summaries of participants’ suggestions for improving the enhanced QA checklist implementation.

### Proposed implementation framework for QA checklists in radiation oncology

All determinants (barriers and facilitators) identified in this study were used to generate an output from the CFIR-ERIC matching tool (Fig 3). We performed this matching with all identified determinants regardless of whether they were classified as barriers or facilitators, for two reasons: determinants classified as facilitators are generally framed as methods to overcome perceived barriers, and the proposed implementation framework should ideally also include strategies that support facilitators. This method has been used in similar studies applying the CFIR-ERIC matching tool [38].

**Fig 3.**
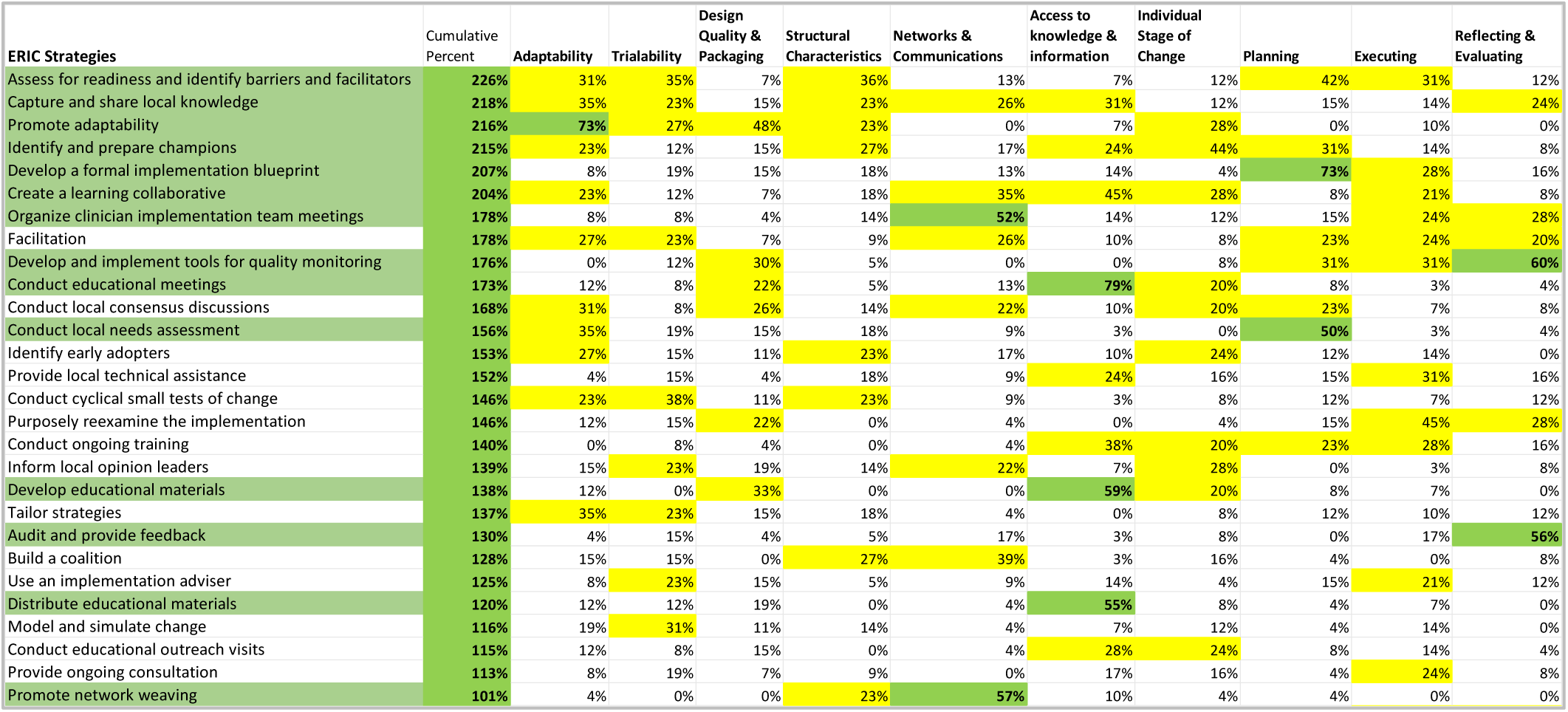
Strategies resulting from the CFIR-ERIC Implementation Strategy Matching Tool. Output of the matching tool showing ERIC strategies matched to the CFIR determinants identified in this study, with the cumulative percentage of expert endorsement.

The top seven strategies with the highest cumulative percentage match with the CFIR determinants were included in the proposed framework, along with the strategies with the highest individual percentage match with CFIR determinants. Thus, we selected 14 evidence-based strategies belonging to 4 clusters from the CFIR-ERIC matching tool for the proposed implementation framework for QA checklists in radiation oncology. A temporal perspective along the three distinct phases of implementation (pre-implementation, implementation, and post-implementation) was added, and these recommendations are specific to radiation oncology settings developing in-house HIT tools such as QA checklists (Fig 4). Our recommendations essentially apply to large academic cancer hospitals where radiation oncology professionals and implementation researchers are involved in implementing QA checklists.

**Fig 4.**
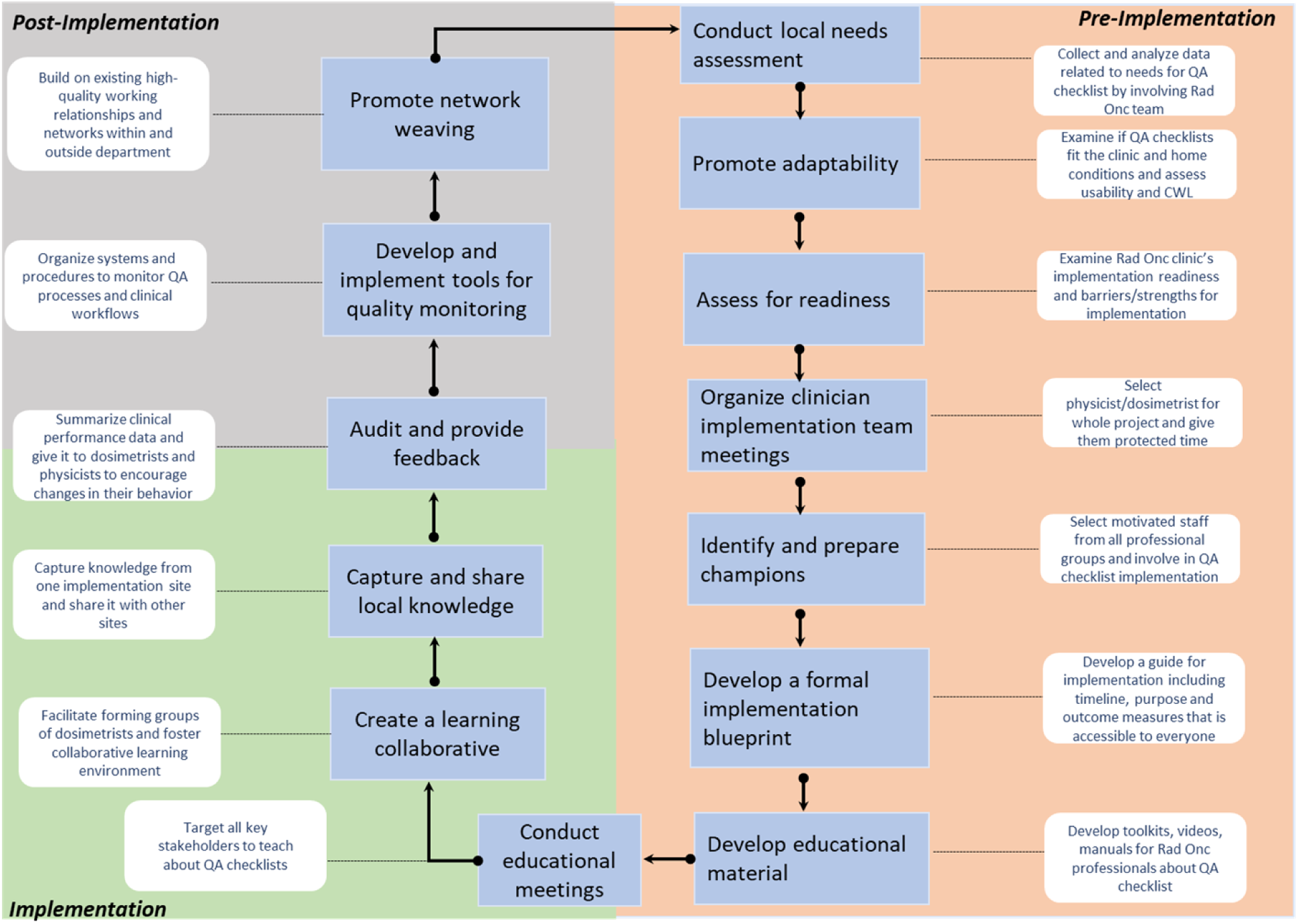
Proposed implementation framework for QA checklists in radiation oncology. Fourteen evidence-based ERIC strategies organized across the pre-implementation, implementation, and post-implementation phases.

In the proposed framework, the pre-implementation phase includes 7 of the 14 strategies, highlighting the importance of key process steps (e.g., assessing local needs) and engaging clinician stakeholders early in the implementation life cycle. During the implementation phase, it is important to focus on creating a learning collaborative based on rich educational meetings and sharing best practices across sites. We recommend applying the ERIC strategy of audit and providing feedback continuously during the entire process and, most definitely, during the implementation phase. The implementation cycle is circular, and some strategies may need to be applied early or repeated at different stages. For instance, promoting network weaving could optimize the implementation context; in our experience, network weaving is more likely when teams achieve project goals during early implementation, which in turn promotes networking with colleagues at outside institutions. We also recognize that, in addition to developing a formal implementation blueprint, it is important to set up an implementation unit with experts on local implementation characteristics and to seek regular feedback from key stakeholder radiation oncology professionals throughout the implementation process. Continuous re-evaluation and quality monitoring are likely to trigger new entries into the implementation strategy, and the implementation cycle will be sustainable only if it is adapted to new user needs.

### Implementation outcomes

#### Acceptability, feasibility, and appropriateness

Table 6 shows the acceptability, feasibility, and appropriateness scores of participants by role. Wilcoxon signed-rank tests showed that acceptability, feasibility, and appropriateness scores during implementation for dosimetrists, physicists, and trainees were statistically significantly higher than at pre-implementation (p<0.05).

**Table 6.** Acceptability, feasibility, and appropriateness scores at pre-implementation and implementation, by participant role.

| Outcome | Dosimetrists: Pre- | Dosimetrists: Impl | p-value | Physicists: Pre- | Physicists: Impl | p-value | Trainees: Pre- | Trainees: Impl | p-value |
| --- | --- | --- | --- | --- | --- | --- | --- | --- | --- |
| Acceptability | 15.25 (0.82) | 18.75 (0.43) | 0.01 | 11.4 (2.48) | 19.4 (0.43) | 0.005 | 8.25 (2.04) | 15.75 (0.43) | 0.004 |
| Feasibility | 15.23 (0.71) | 19.50 (0.87) | 0.006 | 11.20 (0.37) | 18.00 (1.50) | 0.007 | 9.50 (1.50) | 16.25 (1.92) | 0.01 |
| Appropriateness | 16.20 (1.73) | 19.25 (0.83) | 0.03 | 13.20 (1.12) | 18.45 (1.50) | 0.02 | 12.45 (1.58) | 16.25 (1.82) | 0.004 |

Pre- = pre-implementation; Impl = implementation. Values are expressed as mean (standard deviation) of scores on the validated four-item measures (maximum score of 20 per outcome). P-values from Wilcoxon signed-rank tests.

#### Adoption

The adoption rates of the enhanced DQC gradually increased from the first week and reached a 100% adoption rate only in the sixth week of its implementation (Fig 5). Thus, despite statistically significant improvements in acceptability, feasibility, and appropriateness scores in the first weeks of implementation, the adoption rate of the enhanced DQC reached 100% only in week six. The Mann-Whitney U test showed that the mean adoption rates of the enhanced DQC were statistically significantly different from the current DQC (p<0.0001).

**Fig 5.**
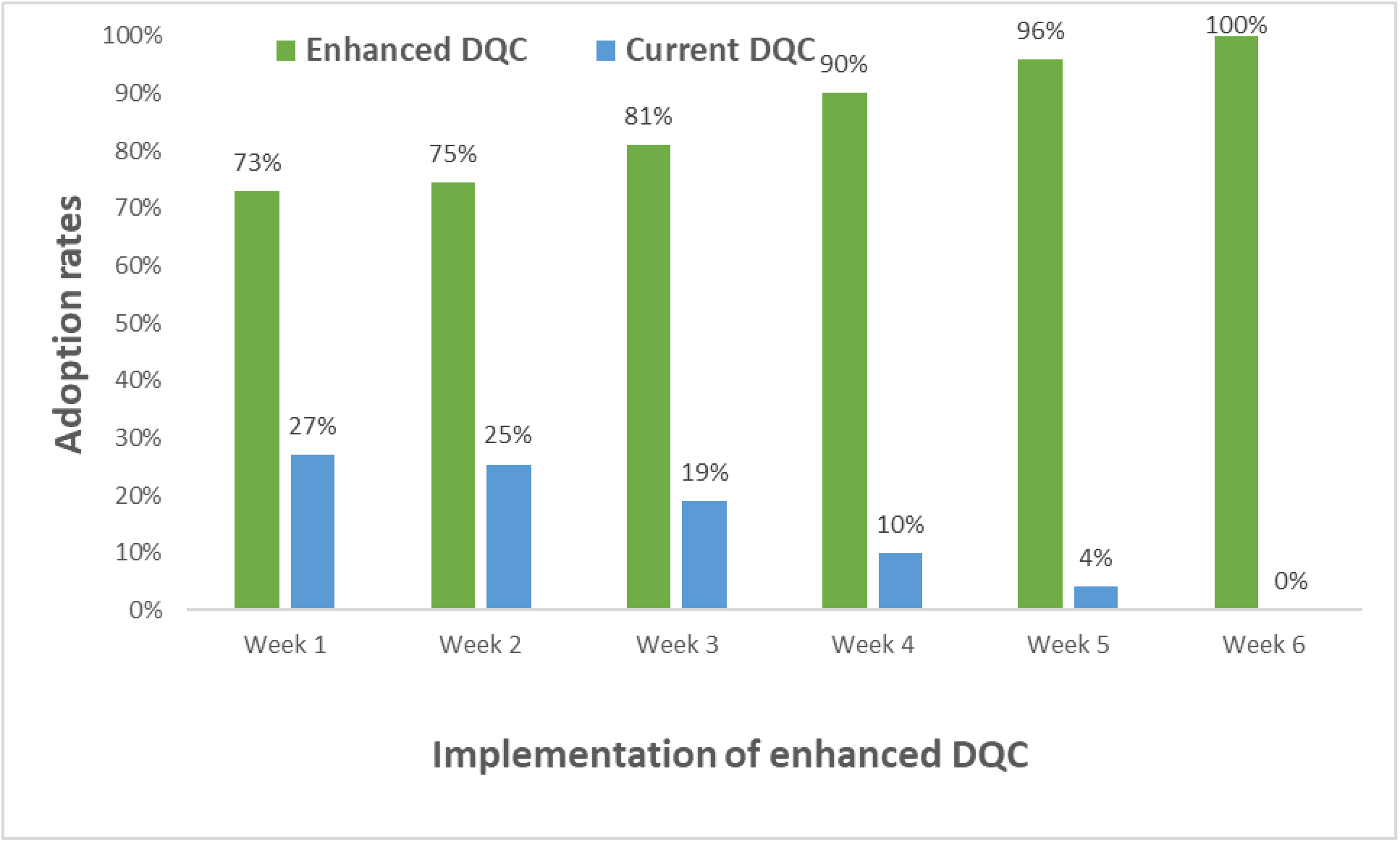
Weekly adoption rates of the enhanced and current DQC during the first six weeks of implementation. Fig 6 presents the adoption rates by participant role for the enhanced and current DQC. The adoption rate for the enhanced DQC was lower than 80% in the first week of implementation for all participants. Because dosimetrists were engaged in all stages of design, development, and implementation, they were the first among all participants to record high adoption rates (>95%), in the third week of implementing the enhanced DQC.

**Fig 6.**
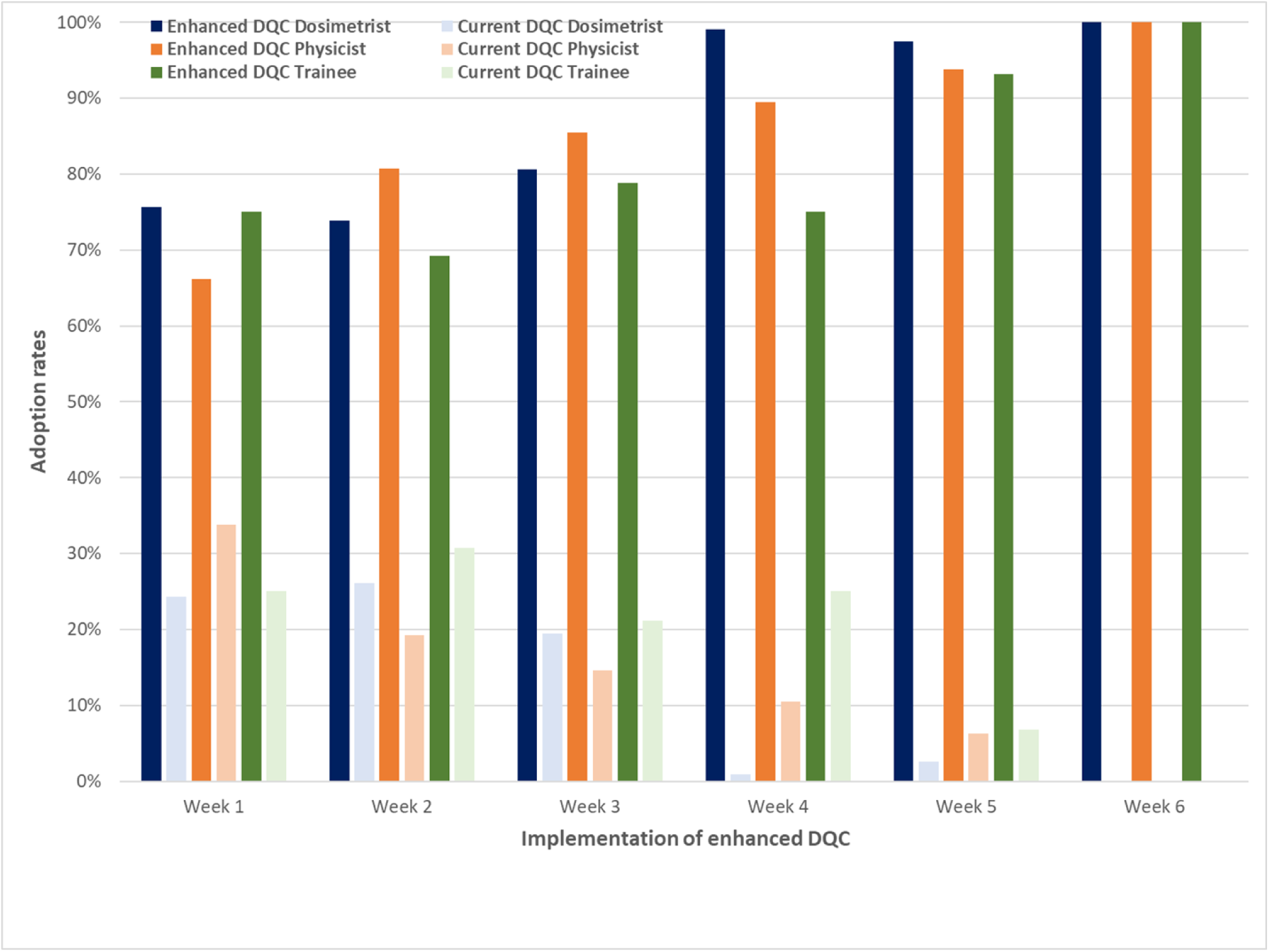
Weekly adoption rates of the enhanced and current DQC by participant role. Reporting determinants, strategies, and outcomes using the IRLM To summarize, we used the IRLM as a reporting tool to present the key determinants (barriers and facilitators), implementation strategies, and outcomes associated with implementing the enhanced DQC in the clinic (Fig 7). This model can now be used to hypothesize relationships between one or more intervening variables, and the nature of those relationships can be formally tested using causal pathway modeling and other path analysis approaches.

**Fig 7.**
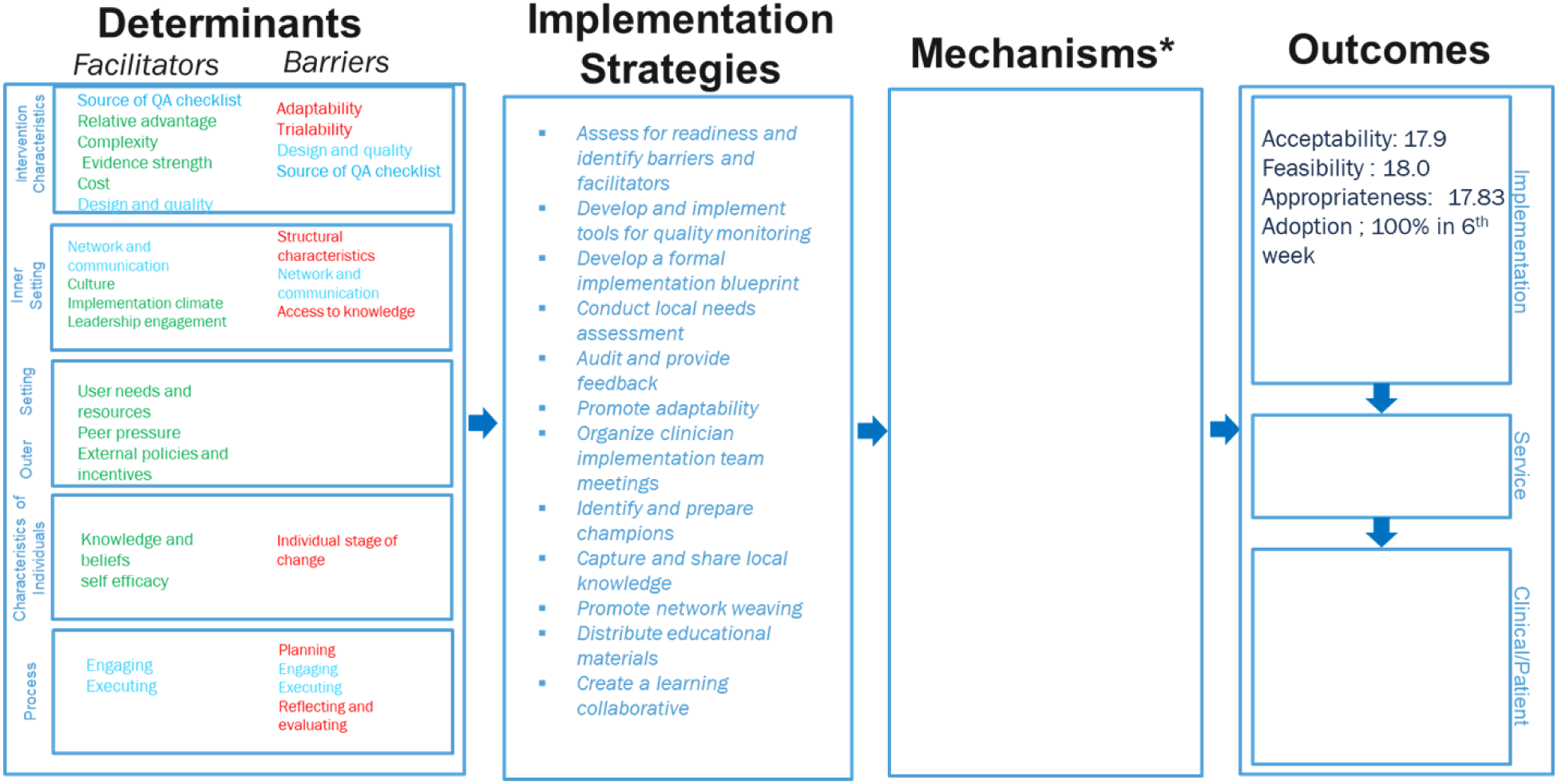
Implementation Research Logic Model showing the determinants, strategies, and outcomes of implementing the enhanced DQC. Determinants with mixed ratings are shown as both facilitators and barriers; mechanisms of action were not considered in this study.

## Discussion

This study identified critical barriers and facilitators for implementing an in-house-built QA checklist in radiation oncology and highlighted 19 ERIC strategies for implementing QA checklists. The proposed implementation framework for in-house QA checklists in radiation oncology includes 14 evidence-based, practical strategies from ERIC clusters to overcome barriers while leveraging facilitators, applicable across the different phases of the implementation life cycle. Overall, this study adds to the growing body of evidence on how the IRLM can effectively report the key determinants, strategies, and outcomes of implementing QA checklists in radiation oncology.

### Determinants and implementation strategies

During pre-implementation and in the general context, it is important to examine end-user needs and lay out the details, aims, and context of the project (conduct local needs assessment; develop a formal implementation blueprint) [58]. Additional strategies (assess for readiness; identify barriers and facilitators) are needed to improve the adaptability of the QA checklist to radiation oncology clinical settings [59]. Previous studies have identified adaptability as a key facilitator of HIT tool implementation in other settings [60,61]. To collaborate creatively with key stakeholders, developers of HIT tools such as QA checklists and human factors researchers should actively participate in the implementation processes, taking advantage of valuable feedback from clinical stakeholders and actively adapting their products based on real-world clinical evidence rather than depending exclusively on controlled laboratory findings [62,63]. Our proposed implementation framework therefore suggests several strategies to enhance the involvement of clinical stakeholders directly (organize clinician implementation team meetings; inform local opinion leaders; identify and prepare champions) and to train and support them (develop educational materials; conduct educational meetings; create a learning collaborative), which resemble strategies proposed for other implementation settings [64]. During the implementation phase, transparent communication of the project’s aims and context (audit and provide feedback) is as critical as effective facilitation for improving user involvement and promoting and sustaining implementation [56].

This study also shows that, using a qualitative approach, we found 19 ERIC strategies organized in 7 clusters to be essential for implementing the enhanced QA checklist in radiation oncology, whereas the CFIR-ERIC matching tool identified 14 strategies belonging to 4 clusters. Using these two distinct approaches, we identified ERIC strategies that could effectively support the implementation of enhanced QA checklists in radiation oncology. Future studies are required to determine the validity and reliability of each method and to differentiate their relative strengths and limitations in identifying optimal strategies for implementing QA checklists in radiation oncology.

### Implementation framework

Implementation science is a relatively young field and has primarily been applied to public health interventions [11]. There are limited implementation science projects in radiation oncology in general and for QA checklists in particular [65]. We recognize that many implementation frameworks have been published, either for specific healthcare settings or as general guidelines [19,24,66–69]. However, few studies have applied implementation science frameworks in radiation oncology settings to propose an implementation framework that includes evidence-based strategies, and explicit guidelines for implementing in-house HIT tools such as QA checklists are completely lacking. The proposed implementation framework fills this gap: it draws on current implementation science research and an interdisciplinary approach specific to radiation oncology settings, and it considers aspects related to implementing QA checklists in academic medical centers. This framework holds promise but requires rigorous future validation in diverse settings.

### Implementation outcomes

The findings of this study highlight the importance of using both subjective and objective measures of implementation outcomes. Acceptability, feasibility, and appropriateness improved significantly among all participants in the first two weeks of implementation. However, adoption rates show that users continued to use the current DQC for over five weeks before completely shifting to the enhanced DQC, suggesting that users continued to rely on the current DQC until they were completely confident in the enhanced DQC. This study also highlights the importance of combining human factors and human-computer interaction measures, such as usability and cognitive workload, with implementation science metrics, such as adoption. In our previous studies, the enhanced DQC demonstrated superior perceived usability, reduced cognitive workload, and improved performance relative to the current DQC [7,10]; the present study shows that favorable usability and early perceptions alone did not guarantee immediate adoption. Notably, several of the strongest facilitators identified here, such as user needs and resources, relative advantage, and confidence in the source of the QA checklist, are plausibly downstream consequences of the participatory co-design and TURF-based design processes used to create the enhanced DQC [8,9], underscoring the continuity between design-stage stakeholder engagement and implementation-stage determinants. Implementation teams must therefore closely monitor the adoption rates of HIT tools and continuously seek feedback from users, even after “go live” in the clinical environment.

### Limitations

This study was conducted at a single academic medical center, and the coding and data analysis were conducted by a single researcher as part of doctoral work. However, the research team and study participants were closely involved with the entire implementation project, including the design and development of the enhanced DQC. Our study therefore provides valuable insights into implementing QA checklists in radiation oncology, highlights important strategies for planning an in-house QA checklist implementation project, identifies potential impediments, and suggests solutions to overcome them.

We derived the implementation framework from the CFIR-ERIC matching tool [38], which was introduced in 2019 and needs further validation and evidence. The method used to identify the evidence-based strategies in this study has been used in various previous studies; nonetheless, future multi-institutional studies in varied settings are required to validate the strategies included in this framework. In addition, mapping strategies to the major barriers might not always lead to the best strategy to tackle a given barrier; we sought to overcome this limitation through discussions within the research team, extensive field research, and analysis of participants’ suggestions.

Finally, we recognize that every radiation oncology department has unique structural and sociotechnical features that could limit the general validity of the derived findings. For instance, our department is a pioneer in patient safety, encourages a multi-disciplinary team approach, and our institution uses tiered huddle systems to encourage the reporting of patient safety events, which may not be the case in other academic medical centers. While we interviewed participants at different stages of implementation, the actual number of interviewees was limited, and there could be potential selection bias among the key stakeholders interviewed throughout the study. This study depicts an implementation project in one radiation oncology clinic, and translation of our findings to other contexts should be done with these specificities in mind. We also recognize the need for further validation and evaluation of our proposed implementation framework in other radiation oncology settings to fully realize its potential for implementing in-house tools such as QA checklists.

## Conclusions

We identified determinants, examined strategies, and assessed outcomes for implementing an enhanced DQC in a radiation oncology clinic, completing a program of research that spanned participatory co-design [8], theory-driven design [9], and human factors evaluation of usability, workload, performance, and patient safety [7,10]. The proposed implementation framework outlines evidence-based strategies that can be applied to implementing in-house-built QA checklists during different phases of the implementation cycle. This framework may facilitate clinicians and implementation advisors in the practical execution of QA checklists in radiation oncology settings.

## Supporting information

**S1 Table.** Original and adapted CFIR constructs mapped to UTAUT constructs and the considerations of Marwaha et al. (2022).

**S1 Appendix.** Adapted CFIR constructs and example interview questions for each CFIR sub-construct.

**S2 Appendix.** Semi-structured interview guide.

**S3 Appendix.** Observation guide.

**S4 Appendix.** Survey instrument (determinants survey administered via Qualtrics).

## Acknowledgments

We thank the dosimetrists, medical physicists, trainees, and software developers of the Department of Radiation Oncology for their participation and support throughout the implementation of the enhanced DQC.

## Author contributions

Conceptualization: K.A., L.M. Data curation: K.A. Formal analysis: K.A. Investigation: K.A. Methodology: K.A., L.M., P.R.M., F.Y., C.M., S.D. Supervision: L.M., S.D. Visualization: K.A. Writing – original draft: K.A. Writing – review and editing: All authors have read and agreed to the published version of the manuscript.

## Data availability

All relevant aggregated data are within the manuscript and its Supporting Information files. The full interview transcripts cannot be shared publicly because they contain potentially identifying information about participants from a single clinical department; de-identified excerpts are available from the corresponding author upon reasonable request for researchers who meet the criteria for access to confidential data.

